# Access to palivizumab among children at high risk of respiratory syncytial virus complications in English hospitals

**DOI:** 10.1101/2021.04.15.21255460

**Authors:** Ania Zylbersztejn, Ofran Almossawi, Nikesh Gudka, Daniel Tompsett, Bianca De Stavola, Joseph F Standing, Rosalind Smyth, Pia Hardelid

## Abstract

**Objectives:** Palivizumab is a monoclonal antibody which can prevent infection with respiratory syncytial virus (RSV). Due to its high cost, it is recommended for high-risk infants only. We aimed to determine the proportion of infants eligible for palivizumab treatment in England who receive at least one dose.

**Methods:** We used the Hospital Treatment Insights database containing hospital admission records linked to hospital pharmacy dispensing data for 43/153 hospitals in England. Infants born between 2010 and 2016 were considered eligible for palivizumab if their medical records indicated chronic lung disease (CLD), congenital heart disease (CHD), or severe immunodeficiency (SCID), and they met additional criteria based on gestational age at birth and age at start of the RSV season (beginning of October). We calculated the proportion of infants who received at least one dose of palivizumab in their first RSV season, and modelled the odds of treatment according to multiple child characteristics using logistic regression models.

**Results:** We identified 3,712 eligible children, of whom 2,479 (67%) had complete information on all risk factors. Palivizumab was prescribed to 832 of eligible children (34%). Being born at <30 weeks’ gestation, aged <6 months at the start of RSV season, and having two or more of CLD, CHD or SCID were associated with higher odds of treatment.

**Conclusion:** In England, palivizumab is not prescribed to the majority of children who are eligible to receive it. Doctors managing these infants might be unfamiliar with the eligibility criteria or are constrained by other considerations, such as cost.

## Introduction

Bronchiolitis due to respiratory syncytial virus (RSV) is a leading cause of morbidity and hospital admissions among young children globally.^1,2^ In England alone, RSV bronchiolitis causes over 40,000 hospital admissions annually^3^ at an estimated cost of £84 million.^4^ Children who were born prematurely or with chronic lung or heart disease (CLD) have an increased risk of bronchiolitis and other RSV-related complications, although around 80% of RSV-related admissions occur among children without a high-risk condition.^5,6^ RSV infection in infancy has also been associated with the development of recurrent wheeze and asthma in later childhood.^7^

There is currently no licensed vaccine against RSV. Only passive immunisation with palivizumab (Synagis®, MedImmune), a humanised monoclonal antibody, is licensed for the prevention of RSV infection. Palivizumab has been found in industry funded trials to lead to a 45% reduction in RSV-related hospital admission rates in children with haemodynamically significant heart disease, and 55% reduction in children born at less than 35 weeks’ gestation or with bronchopulmonary dysplasia.^8,9^ However, palivizumab is a high-cost treatment which requires up to five monthly doses during the RSV season (October to February in the UK). The average cost of a full course of palivizumab treatment for one child has been estimated at £5,000 in the UK.^10^ Due to its high cost, in most countries palivizumab is recommend for a more select group of infants than those included in original clinical trials.^11–14^ In the UK, palivizumab should be administered under expert supervision to preterm infants with CLD or congenital heart disease (CHD), who meet additional strict criteria based on gestational and chronological age, and those with severe combined immunodeficiency (SCID, all criteria are detailed in box 1).^14^ Infants in neonatal units are considered to have low risk of infection and should only begin treatment within 24-48 hours prior to discharge.^14^ In response to the COVID-19 pandemic, the eligibility criteria have recently been expanded to cover all infants with CLD born at ≤34 weeks.^15^

### Box 1 Recommendations for palivizumab use in high-risk children in the UK^14^

A. Infants with bronchopulmonary dysplasia (chronic lung disease of prematurity) who were:
  ‐ Aged <1.5 month at the start of RSV season (October) & born at <34 weeks’ gestation
  ‐ Aged 1.5-3 months at the start of RSV season & born at <32 weeks’ gestation
  ‐ Aged 3-6 months at the start of RSV season & born <28 week’s gestation
  ‐ Aged 6-9 months at the start of RSV season & born at <24 weeks’ gestation
B. Infants who require oxygen at the start of RSV season (preterm or not) aged <1 year at the start of RSV season (October)
C. Infants with haemodynamically significant, acyanotic congenital heart disease who were:
  ‐ Aged <1.5 month & born at <32 weeks’ gestation
  ‐ Aged 1.5-3 months & born at <30 weeks’ gestation
  ‐ Aged 3-6 months & born <26 week’s gestation
D. Infants with congenital heart disease aged <12 months who have significant co-morbidities
E. Children with severe combined immunodeficiency aged <24 months at the start of RSV season (October)
F. *Additionally, palivizumab could be prescribed to infants who are at high risk of complications from RSV based on clinical judgement of other patient circumstances*

Despite palivizumab being recommended to high-risk groups of infants, there is no national dataset in any UK country that allows monitoring the proportion of eligible children that receive palivizumab treatment, and whether access to palivizumab is equitable among eligible English children. The aim of this study was to examine patterns of palivizumab prescribing in England.

## Methods

### Data sources

We developed a study cohort using the Hospital Treatment Insights (HTI) database, which links pharmacy dispensing records for 43 participating hospitals (28% of all hospitals in England) and patient’s hospital records from Hospital Episode Statistics (HES).^16^ Births and prescribing captured in HTI are representative of population-level trends in England (comparison with national statistics is described in appendix 1.1 and 1.3). HTI is maintained by IQVIA (https://www.iqvia.com/).

HES is the national administrative hospital database in England containing details of patient care (inpatient admissions, outpatient appointments and A&E visits) funded by the English NHS since 1^st^ April 1997.^17^ HES covers an estimated 98-99% of all hospital activity and 97% of all births in England.^17^ Recorded data includes diagnoses (coded using the International Classification of Diseases version 10, ICD-10) and procedures (coded using the Office of Population Censuses and Surveys system, OPCS),^17^ administrative details (including dates of admission/discharge), and some patient demographic information (age, sex). Birth admission records include additional information on birth characteristics such as gestational age and mode of delivery. An individual’s hospital contacts can be linked over time in HES using the HESID, a study-specific identifier derived by the data provider, NHS Digital.^18^

Pharmacy dispensing data in HTI are derived from hospital pharmacy systems in the 43 participating hospitals (referred to as HTI hospitals throughout) since 1^st^ January 2010. Recorded details include drug strength, brand, quantity, and dispensing date. Pharmacy dispensing records for drugs ordered for a named patient can be linked to patient records in HES. Data linkage is carried out by NHS Digital (details of the linkage algorithm is described elsewhere^16^).

HTI includes a subset of HES records from the 43 HTI hospitals since 1^st^ January 2010. For individuals with at least one linked pharmacy dispensing record, HTI covers all hospital records (from HTI hospitals and non-HTI hospitals) since 1^st^ January 2005. Examples of hospital admission trajectories captured by HTI are illustrated in Figure 1. HTI does not contain the names of the treating hospitals.

**Figure 1.**
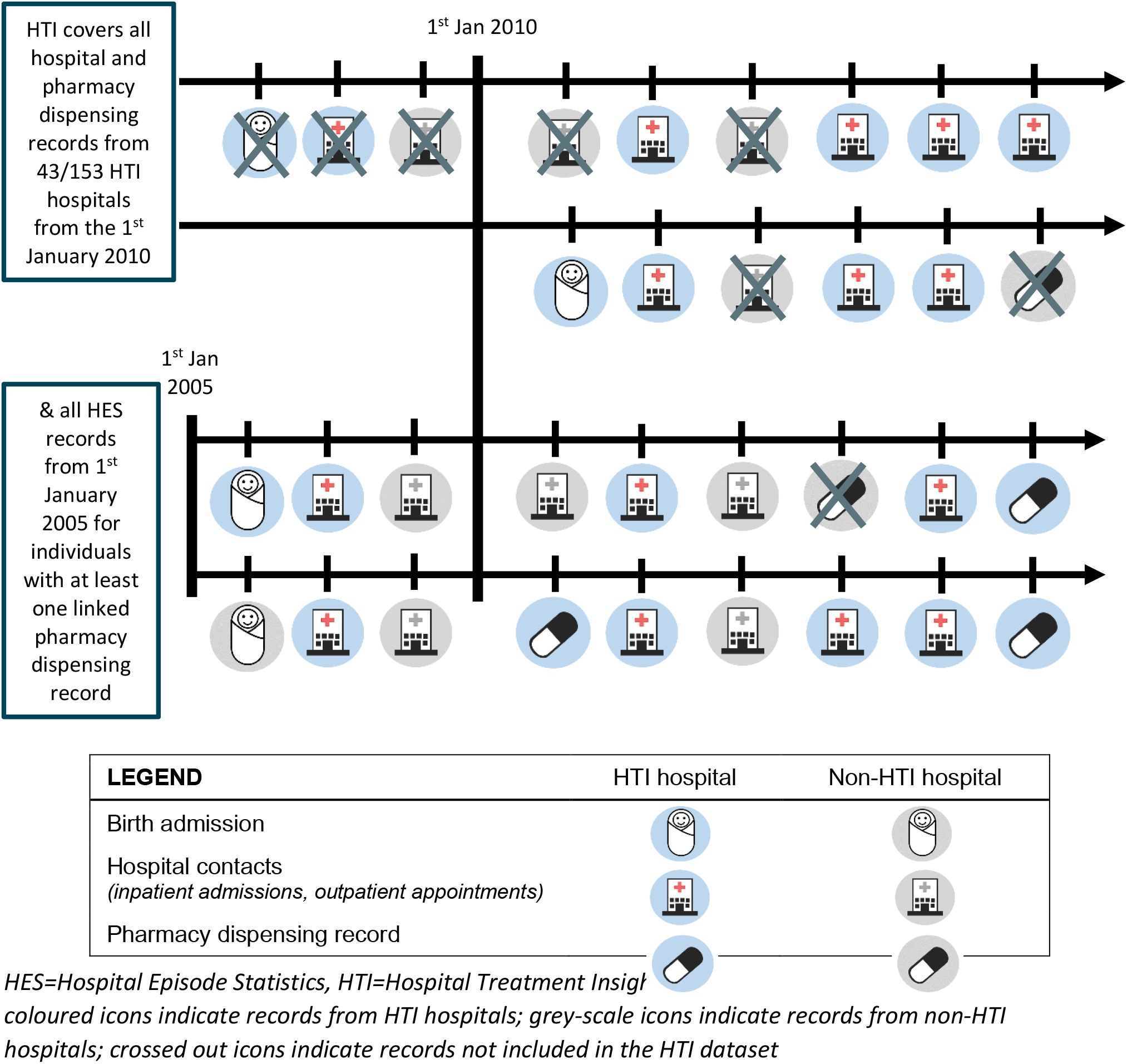
Examples of healthcare trajectories included in HTI

### Study cohort and follow-up

The study cohort included children eligible to receive palivizumab born between 1^st^ January 2010 and 31^st^ December 2016, who were under the care of an HTI hospital. Children were followed up via linkage to HES, from birth until death in hospital, the end of the first RSV season of life or 31^st^ January 2017, whichever occurred first.

We translated the national palivizumab treatment guidelines (summarised in box 1)^14^ into an algorithm for indicating children eligible for palivizumab treatment in HES inpatient admissions records using diagnostic and procedure codes, recorded gestational age and estimated date of birth. First, we derived a cohort of all live births captured in HTI and indicated children with at least one eligible condition recorded as diagnostic or procedure code recorded in the first year of life. Infants under care of non-HTI hospitals (i.e. hospitals that did not contribute pharmacy dispensing data to HTI, indicated by greyscale icons in Figure 1) and those who died before the start of RSV season were excluded. Second, we applied additional exclusion criteria based on gestational and chronological age at start of RSV season. Infants with complete gestational age who did not meet eligibility criteria and infants with missing gestational age for whom eligibility was not possible to indicate were excluded. Lastly, we removed records with missing data on any risk factor of interest for the complete case cohort used for our main analyses. Cohort derivation is described in detail in Appendix 1.2. R scripts used to develop the cohort are available here: https://github.com/UCL-CHIG/Palivizumab

### Outcomes

The main outcome of interest was administration of at least one dose of palivizumab during the first RSV season. This was indicated if an infant had at least one linked pharmacy dispensing record for palivizumab, or OPCS procedure code X86.5 (Respiratory syncytial virus prevention drugs band 1) recorded in inpatient or outpatient records. Since HTI data captures date of dispensing rather than administration of palivizumab, we included any indication of palivizumab between 1^st^ September of the year of birth and 30^th^ April in the following year to allow for early and late recoding of dispensing (4.8% of all pharmacy dispensing occurred in September, March or April). Details of data cleaning are presented in appendix 1.3.

### Risk factors

Information about risk factors was obtained from HES. We obtained information about birth weight (categorised as <1000g, 1000-1499g, 1500-2499g, 2500g+), gestational age (in weeks, categorised as <26, 26-27, 28-29, 30-31, 32-36, 37+), sex, year of birth (as indicator of the first RSV season of eligibility), and age at start of RSV season (1^st^ October) categorised as born before peak RSV season (1^st^ October – 31^st^ December), aged <1.5 months old, 1.5-3 months old, 3-6 months old, 6-9 months old. We used quintile of the Index of Multiple Deprivation (IMD) score as a measure of socioeconomic deprivation. IMD score is a small area indicator measured per 200–1400 households.^17^ The score was allocated by NHS Digital using the child’s postcode recorded at birth or during the earliest hospital admissions in infancy.

We derived a binary complexity indicator to flag whether a child had one or more than one of eligibility conditions (listed in Box 1: A-B for CLD, C-D for CHD, and E for SCID). Presence of other types of comorbidities (congenital anomalies, cancer/blood disorders, chronic infections, metabolic/ endocrine/ digestive/ renal/ genitourinary conditions, musculoskeletal/ skin conditions or other non-specific chronic conditions) was indicated using a code list developed by Hardelid et al^19^ to identify chronic conditions in children. Comorbidities were indicated if any relevant ICD-10 code was recorded at birth, or during any hospital admission in the first year of life.

### Statistical analyses

We evaluated representativeness of HTI by comparing the distribution of birth weight, gestational age, and IMD score for births indicated in HTI compared to national birth statistics published by the Office for National Statistics (ONS).^20,21^

We derived the number and proportion of eligible children and of children who received at least one dose of palivizumab by each risk factor category, overall and in the complete case cohort. Variation in the proportion of children treated between hospitals was examined using a funnel plot.

We used logistic regression models to determine factors associated with increased odds of palivizumab treatment. A priori, we decided to include sex and quintile of IMD score (as confounders) and RSV season (to account for possible changes in dispensing over time). We then added all risk factors that were associated with palivizumab dispensing in univariable analyses to the model (where *p* < 0.1 according to a Chi Squared test; we did not consider birth weight due to correlation with gestational age). Risk factors that did not improve the fit of the multivariable model using likelihood ratio test were excluded.

We additionally fitted a multilevel logistic regression model, allowing for random intercept by hospital to account for possible correlation in prescribing practices within hospitals. We calculated the percentage of variation unaccounted for between hospitals (as captured by the random intercept) due to between-hospital heterogeneity as 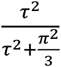 where τ^2^ is the level 2 variance, and 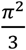 is assumed to be level 1 variance.^22^

### Secondary analyses

We derived the distribution of risk factors among infants who received at least one dose of palivizumab but were not eligible according to the national treatment guidelines or had missing gestational age. We also compared the proportion of infants who died in hospital during RSV season by treatment status.

### Sensitivity analyses

Since non-specific diagnostic codes are often used in HES, our algorithm for indicating palivizumab eligibility used a broad range of diagnostic and procedure codes to ensure that we capture all eligible children, possibly overestimating the number of eligible infants. We therefore repeated all analyses using a stricter definition of eligibility for palivizumab. We included infants who had CLD (eligibility conditions A-B in Box 1), CHD (condition C) or SCID (condition E), who were aged <6 months at the start of RSV season, and whose birth admission finished before the end of RSV season (to indicate and exclude babies likely to be in neonatal intensive care units).

## Results

### Representativeness

We identified 1,395,579 live births in HTI in 2010-2016, covering 34% of all live births in England.^20^ The distributions of birth weight, gestational age and IMD score in births were comparable to published ONS data for England.

### Cohort characteristics

The study cohort comprised 3,712 eligible children (accounting for 0.3% of 1,337,132 infants born in HTI hospitals). 2,479 eligible children (67%) had complete information on all risk factors at birth. Missing data was predominantly driven by gestational age (1,007 children had missing data on GA; 27%) and birth weight (905 children, 24%, Table 1). Missing data rates were comparable with those reported for all births in HTI (Appendix Table 1) and for infants who received palivizumab (Table 1).

**Table 1.**
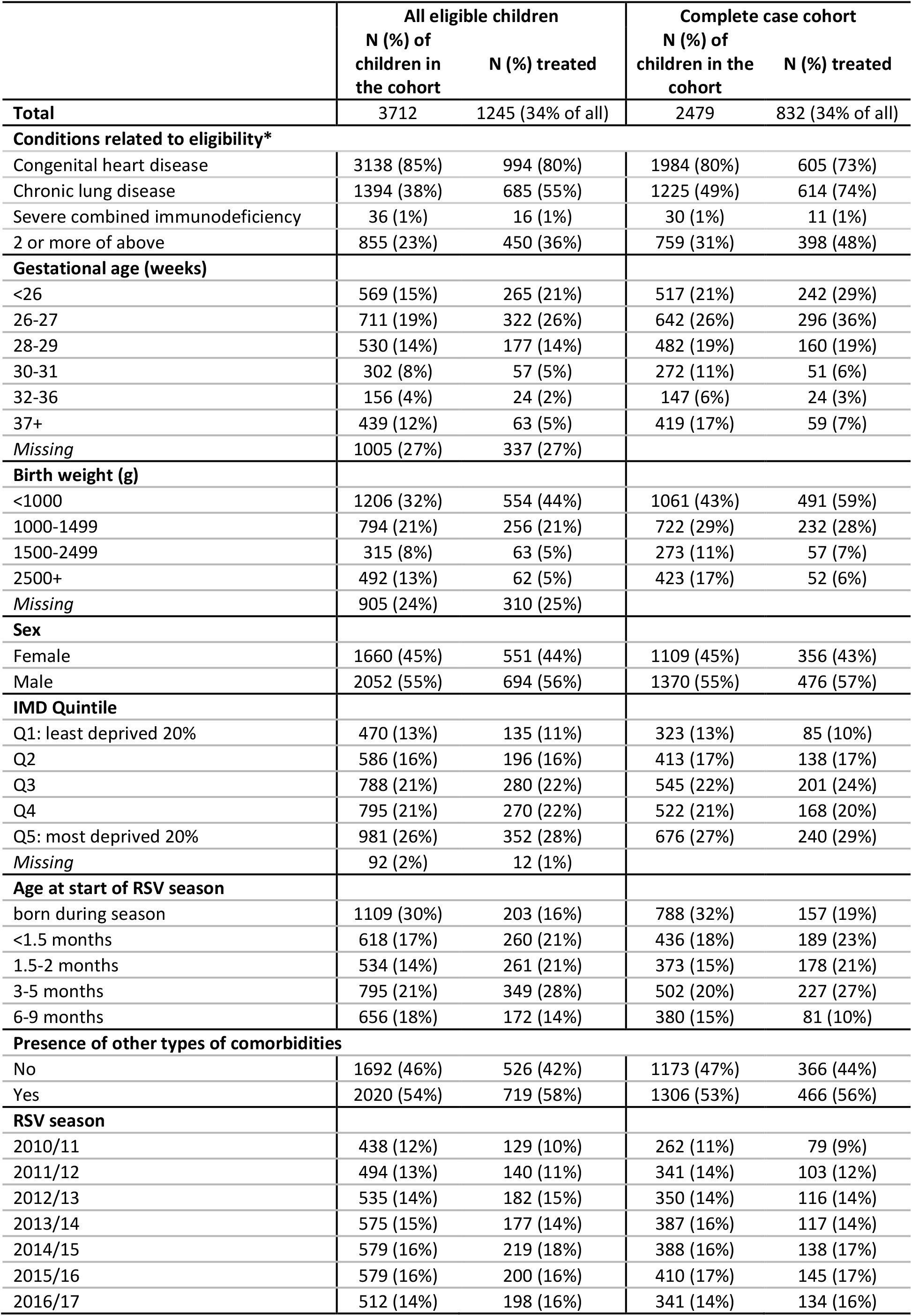
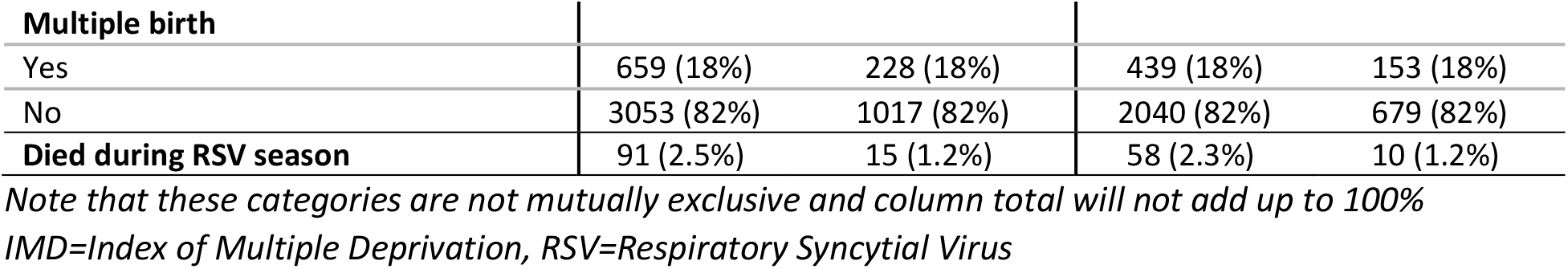
Baseline characteristics of children who were eligible and had complete information on all risk factors, overall and for those who received at least one palivizumab dose in their first RSV season of life

|  | All eligible children |  | Complete case cohort |  |
| --- | --- | --- | --- | --- |
|  | N (%) of children in the cohort | N (%) treated | N (%) of children in the cohort | N (%) treated |
| **Total** | 3712 | 1245 (34% of all) | 2479 | 832 (34% of all) |
| **Conditions related to eligibility\*** |  |  |  |  |
| Congenital heart disease | 3138 (85%) | 994 (80%) | 1984 (80%) | 605 (73%) |
| Chronic lung disease | 1394 (38%) | 685 (55%) | 1225 (49%) | 614 (74%) |
| Severe combined immunodeficiency | 36 (1%) | 16 (1%) | 30 (1%) | 11 (1%) |
| 2 or more of above | 855 (23%) | 450 (36%) | 759 (31%) | 398 (48%) |
| **Gestational age (weeks)** |  |  |  |  |
| <26 | 569 (15%) | 265 (21%) | 517 (21%) | 242 (29%) |
| 26-27 | 711 (19%) | 322 (26%) | 642 (26%) | 296 (36%) |
| 28-29 | 530 (14%) | 177 (14%) | 482 (19%) | 160 (19%) |
| 30-31 | 302 (8%) | 57 (5%) | 272 (11%) | 51 (6%) |
| 32-36 | 156 (4%) | 24 (2%) | 147 (6%) | 24 (3%) |
| 37+ | 439 (12%) | 63 (5%) | 419 (17%) | 59 (7%) |
| Missing | 1005 (27%) | 337 (27%) |  |  |
| **Birth weight (g)** |  |  |  |  |
| <1000 | 1206 (32%) | 554 (44%) | 1061 (43%) | 491 (59%) |
| 1000-1499 | 794 (21%) | 256 (21%) | 722 (29%) | 232 (28%) |
| 1500-2499 | 315 (8%) | 63 (5%) | 273 (11%) | 57 (7%) |
| 2500+ | 492 (13%) | 62 (5%) | 423 (17%) | 52 (6%) |
| Missing | 905 (24%) | 310 (25%) |  |  |
| **Sex** |  |  |  |  |
| Female | 1660 (45%) | 551 (44%) | 1109 (45%) | 356 (43%) |
| Male | 2052 (55%) | 694 (56%) | 1370 (55%) | 476 (57%) |
| **IMD Quintile** |  |  |  |  |
| Q1: least deprived 20% | 470 (13%) | 135 (11%) | 323 (13%) | 85 (10%) |
| Q2 | 586 (16%) | 196 (16%) | 413 (17%) | 138 (17%) |
| Q3 | 788 (21%) | 280 (22%) | 545 (22%) | 201 (24%) |
| Q4 | 795 (21%) | 270 (22%) | 522 (21%) | 168 (20%) |
| Q5: most deprived 20% | 981 (26%) | 352 (28%) | 676 (27%) | 240 (29%) |
| Missing | 92 (2%) | 12 (1%) |  |  |
| **Age at start of RSV season** |  |  |  |  |
| born during season | 1109 (30%) | 203 (16%) | 788 (32%) | 157 (19%) |
| <1.5 months | 618 (17%) | 260 (21%) | 436 (18%) | 189 (23%) |
| 1.5-2 months | 534 (14%) | 261 (21%) | 373 (15%) | 178 (21%) |
| 3-5 months | 795 (21%) | 349 (28%) | 502 (20%) | 227 (27%) |
| 6-9 months | 656 (18%) | 172 (14%) | 380 (15%) | 81 (10%) |
| **Presence of other types of comorbidities** |  |  |  |  |
| No | 1692 (46%) | 526 (42%) | 1173 (47%) | 366 (44%) |
| Yes | 2020 (54%) | 719 (58%) | 1306 (53%) | 466 (56%) |
| **RSV season** |  |  |  |  |
| 2010/11 | 438 (12%) | 129 (10%) | 262 (11%) | 79 (9%) |
| 2011/12 | 494 (13%) | 140 (11%) | 341 (14%) | 103 (12%) |
| 2012/13 | 535 (14%) | 182 (15%) | 350 (14%) | 116 (14%) |
| 2013/14 | 575 (15%) | 177 (14%) | 387 (16%) | 117 (14%) |
| 2014/15 | 579 (16%) | 219 (18%) | 388 (16%) | 138 (17%) |
| 2015/16 | 579 (16%) | 200 (16%) | 410 (17%) | 145 (17%) |
| 2016/17 | 512 (14%) | 198 (16%) | 341 (14%) | 134 (16%) |

| Multiple birth |  |  |  |  |
| --- | --- | --- | --- | --- |
| Yes | 659 (18%) | 228 (18%) | 439 (18%) | 153 (18%) |
| No | 3053 (82%) | 1017 (82%) | 2040 (82%) | 679 (82%) |
| **Died during RSV season** | **91 (2.5%)** | **15 (1.2%)** | **58 (2.3%)** | **10 (1.2%)** |

*Note that these categories are not mutually exclusive and column total will not add up to 100%*
*IMD=Index of Multiple Deprivation, RSV=Respiratory Syncytial Virus*

Of 2,479 eligible children in the complete case cohort, 1,984 (80%) had CHD, 1,225 (49%) had CLD and 30 (1%) had SCID. 759 (31%) had more than one of these conditions. 1,913 (77%) of eligible children were born at <32 weeks’ gestation, 1,783 (72%) had a birth weight <1500g. One third of eligible children (*n*=788) were born in the first half of RSV season (October – December).

### Patterns of palivizumab prescribing in eligible children

1245 (34%) of all eligible children and 832 children in the complete case cohort (34%) received at least one palivizumab dose. This proportion ranged from 0% to 75% between HTI hospitals (Figure 2). Children with CLD, born extremely prematurely (at <28 weeks’ gestation) or with extremely low birth weight (<1000g) were more likely to receive palivizumab (50%, 46% and 46% were treated, respectively). Children born during first half of RSV season (October-December) and aged 6-9 months at the start of RSV season (i.e., born in January-March) were least likely to receive palivizumab (20% and 21% treated respectively compared to 43-48% of children born in other months, Table 1).

**Figure 2.**
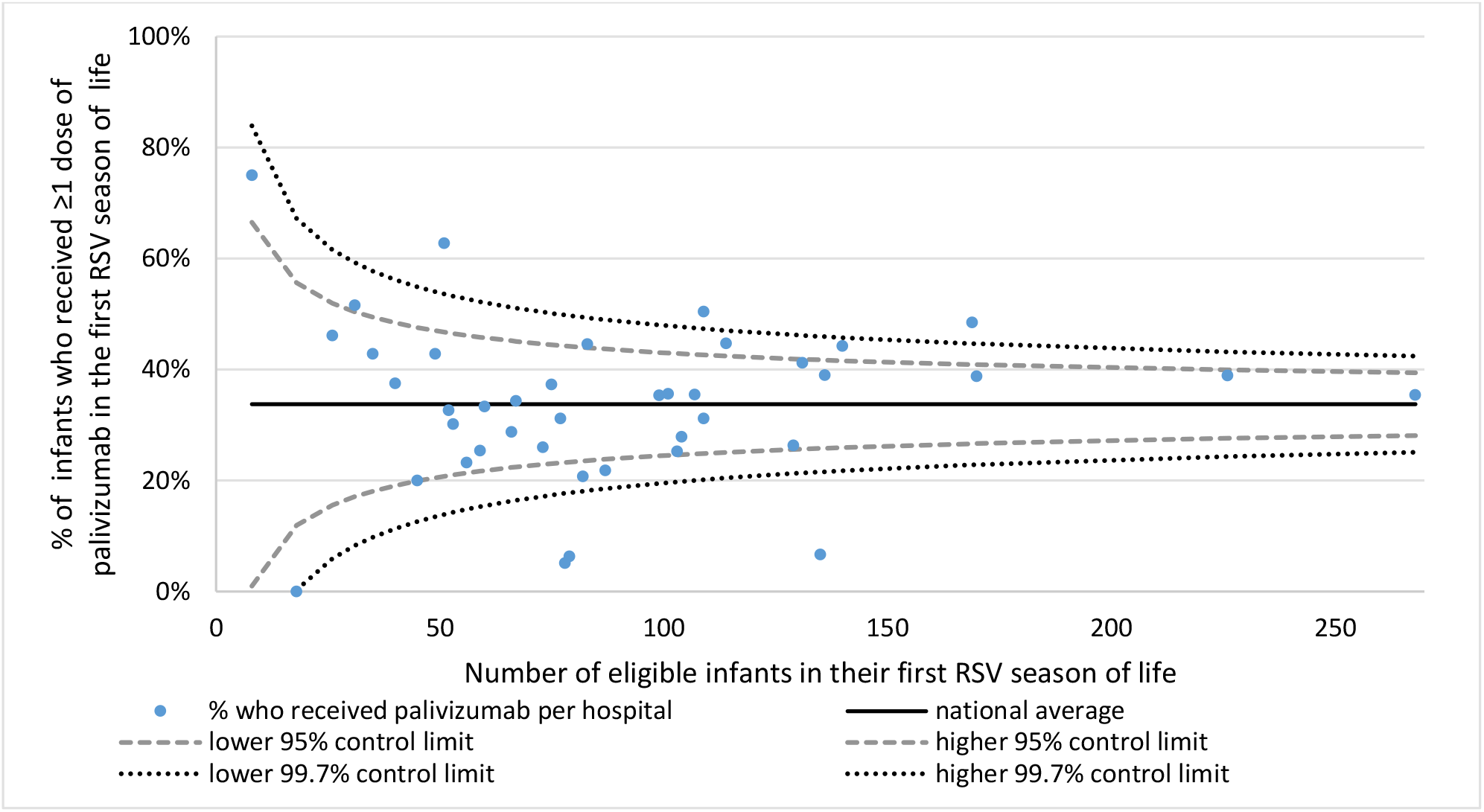
Funnel plot showing variation in palivizumab dispensing by HTI hospital HTI=Hospital Treatment Insights, RSV=respiratory syncytial virus This figure was derived from the number of eligible infants in 41 HTI hospitals (including infants with missing data on any risk factor). We removed two hospitals with <5 eligible infants. Control limits were estimated using normal approximation. 14 out of 41 hospitals (34%) fell outside the 95% control limits (vs 2 that would be expected).

The final model for the odds of palivizumab treatment included sex, RSV season, quintile of IMD score, a measure of complexity, gestational age, an indicator of other chronic conditions, and age at start of RSV season. Babies born at <30 weeks had approximately four times higher adjusted odds of treatment compared to term babies (controlling for the other variables included in the model, Table 2). Infants aged <6 months at the start of RSV season (i.e., born in April-September) had 3-4 times higher adjusted odds for palivizumab compared to babies born before peak RSV season (October-December). Infants with two or more conditions related to eligibility had two times higher adjusted odds of palivizumab treatment. Results were comparable when including a random intercept for hospital (Table 2). Unexplained between-hospital variation, as captured buy the random intercept, contributed to 18% of total observed variation in palivizumab dispensing.

**Table 2.** Results from logistic regression models based on pragmatic eligibility criteria

|  | Fixed effects model<br>Adjusted Odds Ratios<br>(95% Confidence interval) | Random effects model<br>Adjusted Odds Ratios<br>(95% Confidence interval) |
| --- | --- | --- |
| **Complexity (SCID, CHD, CLD)** |  |  |
| 1 condition | 1 | 1 |
| 2 or more conditions | 2.18 (1.75, 2.73) | 2.35 (1.85, 2.98) |
| **Gestational age (weeks)** |  |  |
| <26 | 3.69 (2.57, 5.30) | 4.38 (2.99, 6.40) |
| 26-27 | 4.32 (3.07, 6.09) | 5.14 (3.58, 7.37) |
| 28-29 | 3.10 (2.17, 4.44) | 3.60 (2.47, 5.25) |
| 30-31 | 1.65 (1.07, 2.55) | 1.89 (1.21, 2.98) |
| 32-36 | 1.19 (0.70, 2.02) | 1.27 (0.73, 2.19) |
| 37+ | 1 | 1 |
| **Other chronic conditions** |  |  |
| No | 1 | 1 |
| Yes | 1.34 (1.11, 1.62) | 1.38 (1.13, 1.69) |
| **Age at start of RSV season** |  |  |
| born during season | 1 | 1 |
| <1.5 months | 3.45 (2.62, 4.55) | 3.66 (2.74, 4.89) |
| 1.5-2 months | 3.98 (2.98, 5.31) | 4.62 (3.39, 6.30) |
| 3-5 months | 3.53 (2.69, 4.64) | 4.00 (3.00, 5.34) |
| 6-9 months | 1.62 (1.15, 2.26) | 1.71 (1.20, 2.42) |
| **Sex** |  |  |
| Female | 1 | 1 |
| Male | 1.14 (0.94, 1.37) | 1.10 (0.90, 1.34) |
| **IMD Quintile** |  |  |
| Q1: least deprived 20% | 1 | 1 |
| Q2 | 1.40 (0.99, 2.00) | 1.51 (1.03, 2.20) |
| Q3 | 1.46 (1.05, 2.03) | 1.41 (0.98, 2.02) |
| Q4 | 1.38 (0.98, 1.94) | 1.33 (0.92, 1.92) |
| Q5: most deprived 20% | 1.59 (1.15, 2.19) | 1.50 (1.04, 2.14) |
| **RSV season** |  |  |
| 2010 | 1 | 1 |
| 2011 | 0.95 (0.64, 1.39) | 0.92 (0.61, 1.38) |
| 2012 | 1.13 (0.77, 1.65) | 1.05 (0.70, 1.58) |
| 2013 | 0.91 (0.63, 1.32) | 0.83 (0.56, 1.23) |
| 2014 | 1.35 (0.93, 1.96) | 1.38 (0.93, 2.05) |
| 2015 | 1.41 (0.97, 2.03) | 1.36 (0.92, 2.02) |
| 2016 | 1.45 (0.99, 2.11) | 1.34 (0.90, 2.00) |

CHD=congenital heart disease, CLD=chronic lung disease, IMD=index of multiple deprivation, RSV=respiratory syncytial virus, SCID=severe combined immunodeficiency syndrome

### Secondary analyses

We identified 2,377 children who received at least one dose of palivizumab in their first RSV season of life (Appendix Figure 2): 1245 (52%) children met eligibility criteria, 452 (19%) had an eligible condition (CHD, CLD, SCID) recorded but were missing gestational age, and 680 (29%) did not meet any of the eligibility criteria (listed in box 1).

Infants who did not meet eligibility criteria were older (21% were older than 6 months, compared to 14% of eligible infants), more likely to have normal birth weight (≥2500kg, 29% vs 5% of eligible infants) or be born at term (29% were born at 37+ weeks vs 5% of eligible infants, Appendix Table 4). 207 (30%) of children not meeting eligibility criteria had CHD and 118 (17%) had CLD, but did not meet gestational and chronological age criteria.

58 eligible infants died in hospital during RSV season. 10 received palivizumab (1.2% of 832 treated infants), and 48 did not (2.9% of 1,647 infants who were not treated).

### Sensitivity analyses

We identified 1,801 children who met our stricter eligibility criteria, as defined above, of whom 1,588 (88%) children had complete information on all risk factors. 751 (42%) of these infants overall, and 673 (42%) in the complete case cohort, were prescribed palivizumab. This cohort covered infants with more complex health needs: over 70% of babies meeting these stricter eligibility criteria had either CHD or CLD and 44% had both (Appendix Table 5, compared to 80%, 49% and 31% respectively in the main analysis cohort). Adjusted odds of receiving palivizumab prophylaxis were highest for children aged 1.5-5 months (born in in April-August, Appendix Table 6), born at <28 weeks, and those with a combination of CHD, CLD or SCID.

## Discussion

### Key findings

The majority of children who are eligible for Palivizumab in England are not prescribed it. While 0.3% of children born in England met eligibility criteria for palivizumab, only a third of eligible infants were prescribed palivizumab. Doctors managing these infants are either insufficiently familiar with the guidance or may have felt constrained not to prescribe indicated treatment because of cost. The odds of receiving palivizumab prophylaxis were highest for children aged <6 months old at the start of RSV season (that is, born between April and September), babies born at <30 weeks’ gestation and those with more complex health needs (children with a combination of CHD, CLD or SCID or other chronic conditions). A third of all palivizumab prescriptions were to infants with no indication, and we were not able to determine why they received palivizumab. They might include patients deemed to be at high risk of complications from RSV based on clinical judgment or indicate an inappropriate use of an expensive resource.

### Strengths & limitations

This is the first study to examine variation in access to palivizumab for eligible infants in England. We used a unique source of linked routinely collected admission records and pharmacy dispensing data from multiple hospitals, representative of the population of children in England. Our algorithm identified approximately 0.3% of children to be eligible for palivizumab in England, an estimate comparable to that reported by the UK Joint Committee on Vaccination and Immunisation.^23^ Our R scripts used to develop the cohort are freely available on UCL Child Health Informatics Group’s GitHub repository (https://github.com/UCL-CHIG/Palivizumab) and provide a valuable resource for future work on palivizumab prescribing in England.

Our results could be underestimated due to imperfectly measured denominator population of children eligible for palivizumab. Translating clinical guidelines into clinical codes recorded in electronic health records is not straightforward and we made a number of assumptions (listed in column C, Appendix Table 2). For example, some ICD-10 and OPCS codes are not specific enough to indicate eligibility criteria and we were not able to distinguish between hemodynamically significant congenital heart disease and less severe congenital heart conditions.^13^ It is not possible to measure the duration of, for example, oxygen therapy in hospital admissions records. Our algorithm for indicating eligible children used a broad range of diagnostic and procedure codes to maximise ascertainment of eligible infants as non-specific diagnostic codes are commonly recorded in HES. Palivizumab prescribing was still low, however, in our sensitivity analyses based on a stricter definition of eligibility (42%) and for well-defined conditions such CLD (50%), and these findings are comparable to findings from other studies interationally.^25–27^

Improvements to completeness of recorded birth characteristics in HES are needed. Gestational age, which was used to determine whether an infant was eligible for palivizumab, was missing for nearly 30% of all births and infants with diagnosis of CHD, CLD or SCID. We could not use multiple imputation to take missing data into account in our models, since multiple imputation require all analyses to be run on the same sample size. In this case, the number of eligible children to be included in the models would vary depending on the imputed gestational age values. However, we demonstrated that the distribution of recorded gestational age in HTI was representative of births in England, and the distribution of risk factors among eligible infants with and without any missing data were comparable. Linkage of HES to NHS Birth Notification database (maintained by NHS Digital) could provide complete information on key birth characteristics for all births.

Since HTI only contains pharmacy dispensing data for 43/152 hospitals in England, our study needed to focus on infants under care of these HTI hospitals and exclude infants who might receive palivizumab elsewhere. Full hospital admission history (from HTI and non-HTI hospitals) is required to determine the most likely hospital of care, but this information is only available for individuals with at least one linked pharmacy dispensing record. We therefore excluded infants who did not have at least one linked pharmacy dispensing record (of any kind). Infants eligible for palivizumab have complex health needs and were more likely than other children to have at least one linked pharmacy dispensing record (72% of infants with an eligibility diagnosis had a linked dispensing record compared to 19% of all births in HTI). HES records for all patients (from both HTI and non-HTI hospitals) are required to fully harness the potential of HTI for studies of palivizumab and other medicines prescribed to young children in hospital. Further, our investigation was limited to deaths in hospital. Linkage to national mortality data from the ONS is needed to account for babies who have died in a hospice or at home.

Lastly, we were not able to look at adherence to palivizumab treatment, which requires appropriate number of doses within appropriate time window. There is no consensus on how palivizumab adherence should be defined – previous studies looked at the proportion and number of expected doses that were received, appropriate timing between doses, or a combination of both.^24^ The HTI database covers information about strength and quantity of dispensed drugs, however available information indicates dispensing and not administration. HTI data may not accurately capture the timing of a dose if multiple vials were dispensed at the same time. We also were not able to assess baby’s weight to determine how many vials would be required.

### Interpretation

In this large, population-based study, between a third and 40% of eligible infants were prescribed palivizumab (in main vs sensitivity analyses, respectively). Our findings were comparable to other studies: 47% of 176 eligible infants in Sweden in 2005-2010, 35% of 40 eligible infants in the US in 2000/1 and 41% of 2395 eligible children in Florida received at least one dose of palivizumab;^25–27^ although a study of eligible infants in NICUs in 2003-13 from Western Australia found that only 9.2% of 2679 infants were treated.^11^ During the COVID-19 pandemic, palivizumab treatment has been recommended to all infants with CLD born at ≤34 weeks in the UK.^15^ Our findings suggest that ensuring all eligible infants receive treatment should be prioritised.

Relatively low prescribing rates could reflect variation in practice between hospitals and/or clinicians, and concerns about the cost-effectiveness of an expensive drug requiring multiple injections used in a very high-risk population. Children with CLD were more likely to receive palivizumab than children with CHD (50% vs 30% in the main analyses and 52% and 40% in sensitivity analyses, respectively), in line with findings from other countries.^11,25,27^ Prescribing practices could therefore vary systematically between cardiology and respiratory medicine specialists. We also observed substantial variation in prescribing rates between hospitals. This might partially reflect differences in practice by hospital type (e.g. in secondary vs tertiary hospitals, with/without intensive care facilities on site) which we were not able to assess.

Infants born in the first half of RSV season (October-December) accounted for a third of the cohort, but had the lowest adjusted odds of receiving palivizumab (only 20% were treated compared 43-48% of infants aged 0-5 months in October). The JCVI guidelines allow for the treatment to start during RSV season for eligible infants. This subgroup might include infants in neonatal intensive care units, or infants whose eligibility condition is diagnosed later in the RSV season. Further work is needed to determine why these infants are less frequently considered for palivizumab treatment, and whether this has adverse impact on their health outcomes.

Palivizumab has been shown to lead to a 55% reduction in RSV-related hospital admission rates in infants born at <35 weeks’ gestation,^8^ and 45% reduction in infants with hemodynamically significant congenital heart disease in randomised controlled trials.^9^ In the UK, a much more select group of children with chronic conditions meet eligibility criteria than those included in original clinical trials or in other countries, including Australia, United States and Canada, where studies have been carried out to evaluate real-world effectiveness of palivizumab.^12,28,29^ We found that a lower proportion of treated infants died in hospital during RSV season; however we could not consider out-of-hospital mortality. Our methods for developing a cohort of infants eligible for palivizumab prophylaxis could be used to carry out an emulated target trial of palivizumab effectiveness in preventing RSV-associated admissions and mortality, and to compare long-term outcomes between treated and untreated children.^30^

This study demonstrates the importance of linked hospital pharmacy and admission data for assessing the access to, and real-world effectiveness of, medicines prescribed in hospital. Hospital spending on drugs has been increasing by 12% annually since 2010 in England but our understanding of these trends is limited by lack of data.^31,32^ These cost increases partially reflect rise in use of expensive biological or bioequivalent drugs such as palivizumab, which are likely to be prescribed to a named individual and therefore captured in HTI. HTI could therefore be used to monitor the use of such drugs, provided HES records from all hospitals and linked ONS mortality records are included. More comprehensive data are needed to explore the use of other medications which may be dispensed in blocks to a ward and not to a person, including antibiotics.^16^

## Conclusions

We found that only one third of eligible children have received at least one dose of palivizumab. Doctors managing these infants are either insufficiently familiar with the guidance or may feel constrained not to prescribe indicated treatment because of cost. Further work is needed to establish the effectiveness of palivizumab in the UK given the specific high-risk population eligible to receive it.

## Data Availability

The HTI is maintained by IQVIA (https://www.iqvia.com/). Copyright (c) 2019, re-used with the permission of The Health & Social Care Information Centre. All rights reserved. Copyright (c) 2021, re-used with the permission of IQVIA. All rights reserved.
Authors do not have permission to share patient-level HTI data. Qualified researchers can request access to the data from IQVIA (contact Tanith Hjelmbjerg). All study protocols are subject to review by an Independent Scientific and Ethical Advisory Committee (ISEAC).

## Footnotes

## Acknowledgments

We would like to thank Hassy Dattani and Tanith Hjelmbjerg for their help with accessing the HTI database. We would also like to thank Matthew Jay for his help with R.

This work uses data provided by patients and collected by the NHS as part of their care and support.

## Contributions

PH conceptualised the study. AZ cleaned and analysed the data. AZ and PH wrote the first draft of the manuscript. All authors contributed to writing and finalising the manuscript.

## Data sharing statement

The HTI is maintained by IQVIA (https://www.iqvia.com/). Copyright (c) 2019, re-used with the permission of The Health & Social Care Information Centre. All rights reserved. Copyright (c) 2021, re-used with the permission of IQVIA. All rights reserved

Authors do not have permission to share patient-level HTI data. Qualified researchers can request access to the data from IQVIA (contact Tanith Hjelmbjerg). All study protocols are subject to review by an Independent Scientific and Ethical Advisory Committee (ISEAC).

## Ethics approval

This study protocol was approved by Independent Scientific and Ethical Advisory Committee (ISEAC), protocol number HTI_17ISEAC_001.

