## Supplemental Materials for "Access to palivizumab among children at high risk of respiratory syncytial virus complications in English hospitals"

### Appendix 1 – deriving the study cohort

Appendix 1 describes methods for developing a cohort of children eligible for palivizumab or treated using Hospital Treatment Insights (HTI). R scripts used to develop the cohort are available here: https://github.com/UCL-CHIG/Palivizumab

#### 1.1 Developing a birth cohort in HTI

We first developed a cohort of births between 2010 and 2016 captured in HTI to evaluate representativeness of the data compared to the general population of infants in England. We applied algorithms for developing a birth cohort using HES and data cleaning rules described in detail elsewhere.[1] In brief, we extracted all HES hospital admissions between 1st January 2010 and 31st December 2016 where age at admission was <7 days. We excluded stillbirths, indicated multiple births and derived one record per birth. We identified 1,395,579 live births in HTI in 2010-2016, covering 34% of all live births in England.[2]

We cleaned information on birth weight, gestational age and quintile of Index of Multiple Deprivation (IMD) score using methods published elsewhere.[1] The distribution of live births by birth weight, gestational age and quintile of IMD score was comparable to the distribution among all births in England from national birth statistics published by the Office for National Statistics (Appendix Table 1).[2,3] Rates of missing data were high in HES/HTI as reported previously: 271,600 (20%) of births had missing birth weight, 395,540 (28%) had missing gestational age and 681,898 (49%) had missing IMD score (Appendix Table 3).[1]

*Appendix Table 1 – Distribution of birth characteristics in the birth cohort derived from the HTI database vs the gold standard from ONS*

| | National birth registration data for England (ONS) [2,3] | | | All births in HTI data | | |
|---|---|---|---|---|---|---|
| | N | % of all | % excluding records with missing data | N | % of all | % excluding records with missing data |
| **Birth weight (g)*** | | | | | | |
| Total | 4,964,620 | | | 1,395,579 | | |
| <1000 | 24,535 | 0.5% | 0.5% | 5,344 | 0.4% | 0.5% |
| 1000-1499 | 29,733 | 0.6% | 0.6% | 7,894 | 0.6% | 0.7% |
| 1500-1999 | 67,268 | 1.4% | 1.4% | 15,728 | 1.1% | 1.4% |
| 2000-2499 | 225,182 | 4.5% | 4.6% | 50,418 | 3.6% | 4.5% |
| 2500-2999 | 806,327 | 16.2% | 16.4% | 184,848 | 13.2% | 16.4% |
| 3000-3499 | 1,751,170 | 35.3% | 35.7% | 402,966 | 28.9% | 35.9% |
| 3500-3999 | 1,449,165 | 29.2% | 29.5% | 333,581 | 23.9% | 29.7% |
| 4000+ | 551,882 | 11.1% | 11.3% | 123,200 | 8.8% | 11.0% |
| *Missing* | 59,358 | 1.2% | | 271,600 | 19.5% | |
| **Gestational age (weeks)** | | | | | | |
| Total | 4,722,941 | | | 1,395,579 | | |
| <24 | 4,963 | 0.1% | 0.1% | 548 | 0.0% | 0.1% |
| 24-27 | 15,665 | 0.3% | 0.3% | 3,935 | 0.3% | 0.4% |
| 28-31 | 37,284 | 0.8% | 0.8% | 8,398 | 0.6% | 0.8% |
| 32-36 | 290,884 | 6.2% | 6.2% | 59,802 | 4.3% | 6.0% |
| 37-41 | 4,180,900 | 88.5% | 89.2% | 890,420 | 63.8% | 89.0% |
| 42+ | 156,438 | 3.3% | 3.3% | 36,936 | 2.6% | 3.7% |
| *Missing* | 36,807 | 0.8% | | 395,540 | 28.3% | |
| **IMD scores** | | | | | | |
| Total | 4,722,941 | | | 1,395,579 | | |
| Q1: least deprived 20% | 698,158 | 14.8% | *NA* | 108,628 | 7.8% | 15.2% |
| Q2 | 785,027 | 16.6% | *NA* | 120,944 | 8.7% | 16.9% |
| Q3 | 889,084 | 18.8% | *NA* | 154,859 | 11.1% | 21.7% |
| Q4 | 1,072,623 | 22.7% | *NA* | 163,473 | 11.7% | 22.9% |
| Q5: most deprived 20% | 1,278,049 | 27.1% | *NA* | 165,777 | 11.9% | 23.2% |
| *Missing* | 0 | 0% | *NA* | 681,898 | 48.9% | |

*HTI=Hospital Treatment Insights, IMD=Index of Multiple Deprivation, NA=not applicable, ONS=Office for National Statistics, RSV=respiratory syncytial virus*

**Aggregate data by birth weight category combines live births in England and Wales*

#### 1.2 Developing a cohort of eligible children

Cohort derivation is presented in Appendix Figure 1. The steps were as follows:

**Step 1:** identify all HES birth records captured in HTI (as described in Appendix 1.1)

**Step 2:** indicate potentially eligible children if at least one relevant diagnostic or procedure code were recorded during a hospital admission in the first year of life.

We translated the national palivizumab treatment guidelines (as stated in the 2015 revision of chapter 27a of the Green Book, described in column B of Appendix Table 2)[4] into an algorithm based on diagnostic codes (the International Classification of Diseases version 10 codes, ICD-10), procedure codes (the Office of Population Censuses and Surveys [OPCS] codes), recorded gestational age and estimated time of birth. Eligibility criteria used in step 2 are listed in column C of Appendix Table 2.

**Step 3**: exclude babies who died in hospital before the start of RSV season (1st October each calendar year).

**Step 4:** exclude infants without linked pharmacy dispensing record of any kind

To ensure the cohort does not cover eligible infants who might have received palivizumab treatment in non-HTI hospitals, we needed to exclude infants who were likely to be under the care of non-HTI hospitals (see step 5). Full hospital admission history (from HTI and non-HTI hospitals) is required to determine the most likely hospital of care. This information was only available for infants with at least one pharmacy dispensing record (of any kind). We therefore excluded infants with no linked pharmacy dispensing records (step 4 in Appendix Figure 1). 72% of potentially eligible infants and 19% of all births in HTI had at least one linked pharmacy dispensing record.

**Step 5:** exclude infants likely to be under care of non-HTI hospitals

We extracted hospital in-patient and outpatient records from HTI and derived summaries of hospital contacts in the first year of life for each child as follows:

- **HES inpatient admission records**:
  Episodes of hospital admitted patient care were linked into admissions. Hospital admission was defined as a continuous period in hospital; admissions and transfers on the same day were treated as part of the same hospital admission.[5] We kept one record per admission and we summed the number of inpatient admissions according to each hospital (non-HTI trusts were classified into one category: “other”). For each child, we retained information about the two hospitals where the child had the highest number of admissions (represented by columns 2 and 3 of Appendix Table 3).

- **HES outpatient appointment records:**
  We removed appointments marked as unattended, and de-duplicated appointments according to appointment date, treatment specialty, and hospital. We then summed the number of appointments per hospital per child. For each child, information about two hospitals with the highest number of appointments was retained (represented by columns 4 and 5 of Appendix Table 3).

The most likely hospital of care was indicated based on hierarchal set of rules summarised in Appendix Table 3. Infants receiving most of their care in non-HTI hospitals were excluded from the analyses.

**Step 6:** select one baby per multiple birth at random (as outcomes among multiples are likely be correlated)

**Step 7:** apply additional eligibility criteria based on gestational and chronological age

We applied additional gestational age and chronological age eligibility criteria (listed in column D in Appendix Table 2). Infants with complete gestational age who did not meet these eligibility criteria (step 7a) and infants with missing gestational age for whom eligibility was not possible to indicate (step 7b) were excluded.

**Step 8:** exclude records with missing data on any risk factor of interest for the complete case analyses.

*Appendix Figure 1 – Flow chart detailing cohort derivation*

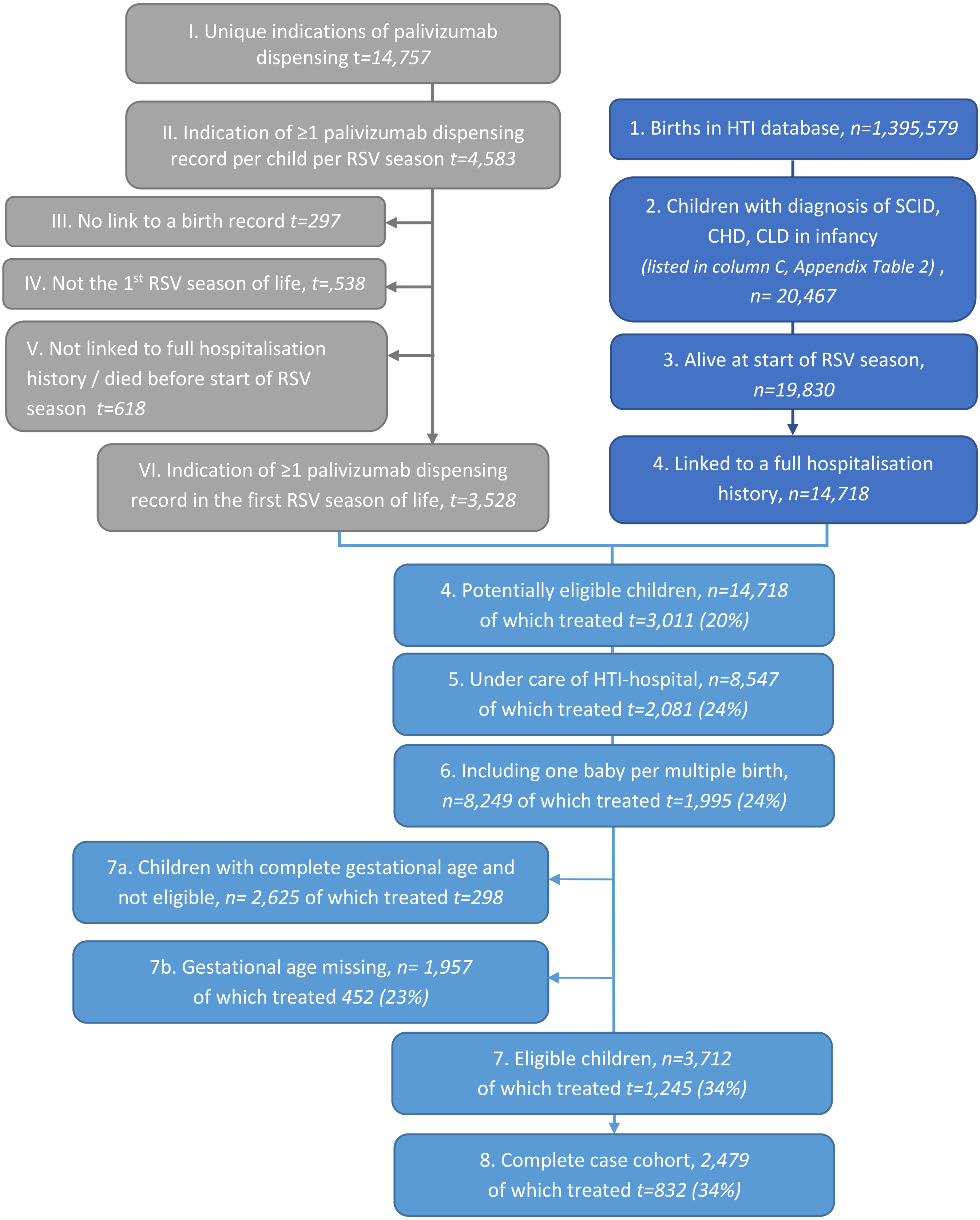


*CHD=Congenital heart disease, CLD=chronic lung disease, HTI=Hospital Treatment Insights, RSV= respiratory syncytial virus, SCID=severe combined immunodeficiency disorder*

*Appendix Table 2 – Eligibility criteria for palivizumab treatment in England according to the Green Book, and an algorithm for identifying children eligible for palivizumab treatment in electronic health records*

| **A. Category** | | **B. Eligibility criteria according to chapter 27a of the Green Book, 2015 edition**[4] | **C. Step 1 – identify infants potentially eligible for palivizumab treatment** | **D. Step 2 – finalise the cohort** |
|---|---|---|---|---|
| **1. Chronic lung disease (CLD)** | 1a | Preterm infants who have moderate or severe bronchopulmonary dysplasia (BPD) at the chronological ages at the start of the RSV season and gestational ages as follows:<br>- Aged <1.5 month & born at <34 weeks' gestation<br>- Aged 1.5-3 months & born at <32 weeks' gestation<br>- Aged 3-6 months & born <28 week's gestation<br>- Aged 6-9 months & born at <24 weeks' gestation<br><br>Moderate or severe BPD is defined as *"preterm infants with compatible x-ray changes who continue to receive supplemental oxygen or respiratory support at 36 weeks post-menstrual age."* | We indicated children with ICD-10 diagnostic code P27 **"***chronic respiratory disease originating in the neonatal period*" recorded anywhere in baby's record at age <9 months. | We excluded children with missing gestational age<br><br>We excluded children who had complete gestational age and did not meet gestational and chronological age criteria (according to age on 1st October) listed in column B. |
| | 1b | Infants with respiratory diseases who are not necessarily pre-term but who remain in oxygen at the start of the RSV season. These infants may include those with conditions including:<br>- pulmonary hypoplasia due to congenital diaphragmatic hernia<br>- other congenital lung abnormalities (sometimes also involving congenital heart disease or lung malformation)<br>- interstitial lung disease<br>- those receiving long term ventilation at the onset of the season | We indicated children with ICD-10 diagnostic codes:<br>- Q33 "Congenital malformations of lung (including hypoplasia)"<br>- Q79.0 "Congenital diaphragmatic hernia"<br>- Q79.1 "Other congenital malformations of diaphragm"<br>- J80 "Adult respiratory distress syndrome"<br>- J81 "Pulmonary oedema"<br>- J82 "Pulmonary eosinophilia, not elsewhere classified"<br>- J84 "Other interstitial pulmonary diseases"<br><br>We indicated children with OPCS procedure codes:<br>- E871 **"**Home oxygen support"<br>- E872 "Long term oxygen assessment"<br><br>We included only infants who had any of the above codes recorded during hospital admissions in October & at age <12 months | *No step 2* |

| | | | | |
|---|---|---|---|---|
| **2. Congenital Heart Disease (CHD)** | 2a | Preterm infants with haemodynamically significant, acyanotic CHD at the chronological ages at the start of the RSV season and gestational ages at birth:<br>- Aged <1.5 month & born at <32 weeks' gestation<br>- Aged 1.5-3 months & born at <30 weeks' gestation<br>- Aged 3-6 months & born <26 week's gestation | We indicated infants aged <6 months with ICD-10 diagnostic codes:<br>- Q20 "Congenital malformations of cardiac chambers and connections"<br>- Q21 "Congenital malformations of cardiac septa"<br>- Q22 "Congenital malformations of pulmonary and tricuspid valves"<br>- Q23 "Congenital malformations of aortic and mitral valves"<br>- Q24 "Other congenital malformations of heart"<br>- Q25 "Congenital malformations of great arteries"<br>- Q26 "Congenital malformations of great veins"<br><br>Indicate children aged <6 months with OPCS procedure codes:<br>- Chapter K (K01-K99) "Heart"<br>- Selected codes from chapter L "Arteries and veins" (L01-L29, L65-L83) | We excluded children with missing gestational age<br><br>We excluded children who had complete gestational age and did not meet gestational and chronological age criteria (according to age on 1st October) listed in column B. |
| | 2b | Infants with cyanotic or acyanotic CHD plus significant co-morbidities, particularly if multiple organ systems are involved | We indicated infants with diagnostic and procedure codes listed above (2a), and a neurological condition from Hardelid code list[6] recorded during any hospital admission at age <12 months. | *No step 2* |
| **3. Severe combined immunodeficiency (SCID) syndrome** | | Children less than 24 months of age with SCID - the most severe form of inherited deficiency of immunity, who are unable to mount either T-cell responses or produce antibody against infectious agents – until immune reconstituted. | We indicated children aged <24 with ICD-10 diagnostic codes:<br>- D81 "Combined immunodeficiencies"<br>- D82: "Immunodeficiency associated with other major defects"<br><br>We used the latest discharge date for admission with any of these codes as time when immunity reconstituted. Children were eligible if they were aged <12 months (as we focus on the first RSV season of life) and this date was after start of RSV season. | *No step 2* |

*BPD: bronchopulmonary dysplasia, CHD: congenital heart disease, CLD: chronic lung disease, ICD-10: International Classification of Diseases version 10, OCPS: the Office of Population Censuses and Surveys system, RSV: respiratory syncytial virus, SCID: severe combined immunodeficiency syndrome*

*Appendix Table 3 – Algorithm for identifying the most likely hospital of care per child based on their HES inpatient and outpatient records*

| Rule nr | **Hosp_APC_1:** Hospital with the highest number of inpatient admissions | **Hosp_APC_2:** Hospital with the 2nd highest number of inpatient admissions | **Hosp_OP_1:** Hospital with the highest number of outpatient appointments | **Hosp_OP_2:** Hospital with the 2nd highest number of outpatient appointments | Selected most likely hospital of care |
|---|---|---|---|---|---|
| 1 | Agree | Not used | Agree | Not used | Hosp_APC_1 =Hosp_OP_1 |
| 2 | no recorded admissions | Not used | Complete | Not used | Hosp_OP_1 |
| 3 | Complete | Not used | no recorded appointments | Not used | Hosp_APC_1 |
| 4 | Not used | Agree | Agree | Not used | Hosp_APC_2 =Hosp_OP_1 |
| 5 | Agree | Not used | Not used | Agree | Hosp_APC_1 =Hosp_OP_2 |
| 6 | Not used | Agree | Not used | Agree | Hosp_APC_2 =Hosp_OP_2 |
| 7 | No agreement | No agreement or no 2nd hospital | No agreement | No agreement or no 2nd hospital | "other" |

*HES: Hospital Episode Statistics*

#### 1.3 Deriving indication of palivizumab treatment from HTI database

##### 1.3.1 Cleaning and validating palivizumab dispensing records

Data cleaning for palivizumab dispensing records is illustrated in the top part of Appendix Figure 2. We identified 16,692 palivizumab dispensing records between 1st September 2010 and 31st January 2017. We removed 1,877 (11%) of dispensing records which were not linked to an individual (these could be dispensed as e.g. ward stock or they could be missed links between HES and dispensing data), and 2,353 (14%) dispensing records with date +/-2 days apart. This resulted in 12,462 records indicated in HTI data.

We additionally looked for indication of palivizumab based on procedure code X86.5 "Respiratory syncytial virus prevention drugs band 1". We identified 2,553 such codes in inpatient records and 1,397 codes in outpatient records. We linked records indicating palivizumab from HTI, HES inpatient and outpatient data, deduplicated records with dispensing date within +/-2 days. HTI data (enhanced with information from HES) included 14,795 records of palivizumab dispensing.

*Appendix Figure 2– Flow chart illustrating data cleaning for palivizumab prescription records*

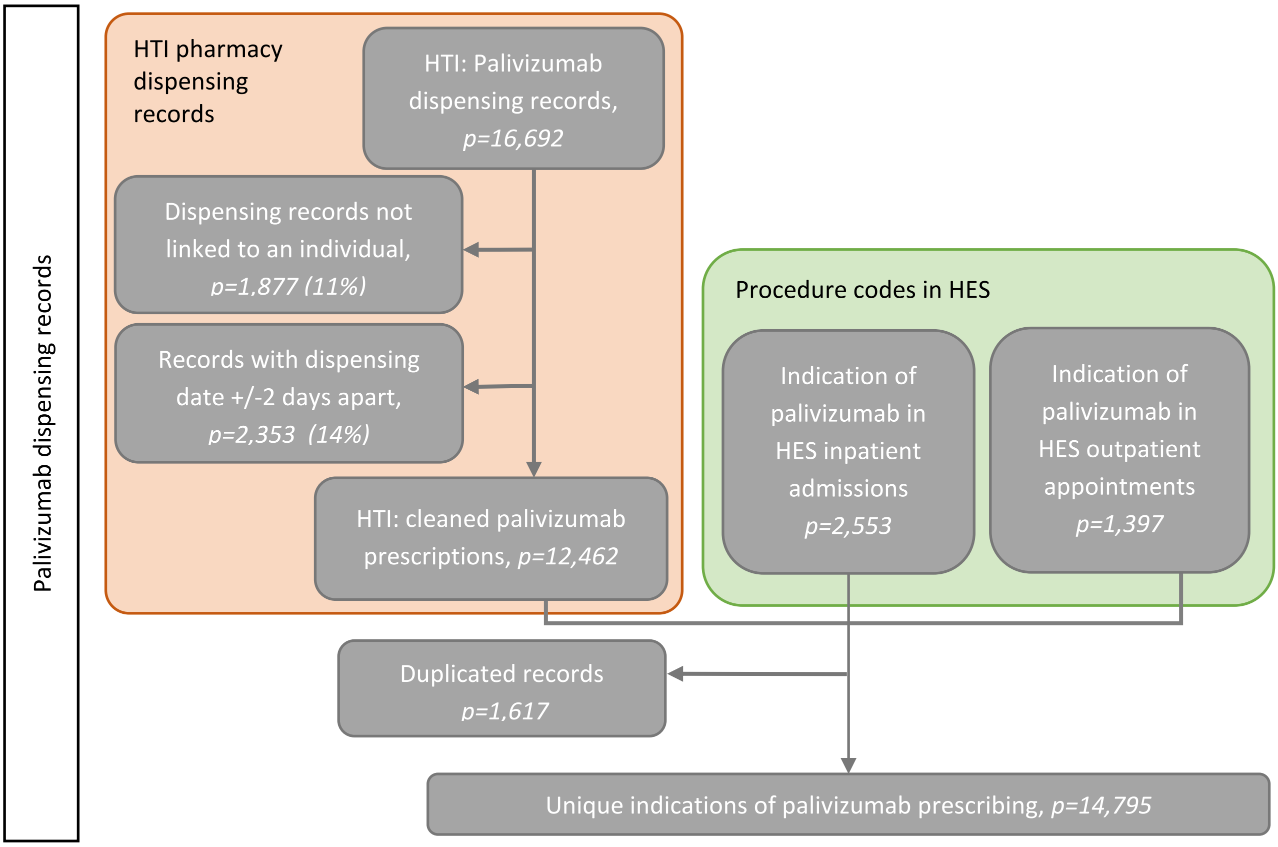


*HES=Hospital Episode Statistics, HTI=Hospital Treatment Insights, RSV=respiratory syncytial virus*

To evaluate representativeness of this data, we compared the distribution of palivizumab dispensing in HTI with nationally representative aggregate hospital dispensing data from the Hospital Pharmacy Audit Index (HPAI) database, also held by IQVIA, by month and year of birth.[7] HTI data covered 14,785 of the 90,333 palivizumab dispensing records indicated in HPAI (16%). This is lower than expected (HTI captures approximately 30% of hospitals in England). Some of the HTI-hospitals may not have paediatric services where palivizumab would be likely to be prescribed. We were not able to explore this further as hospitals are pseudonymised in the HTI database.

Seasonal patterns of palivizumab dispensing were comparable between the two sources (Appendix Figure 3). The number of palivizumab dispensing records in HTI has increased over time, unlike in HPAI. This likely reflected improved linkage between HTI and HES (the proportion of HTI records with no linked HESID declined over time, data not shown).

*Appendix Figure 3 – Number of prescriptions from HTI database compared to HPAI[7]*

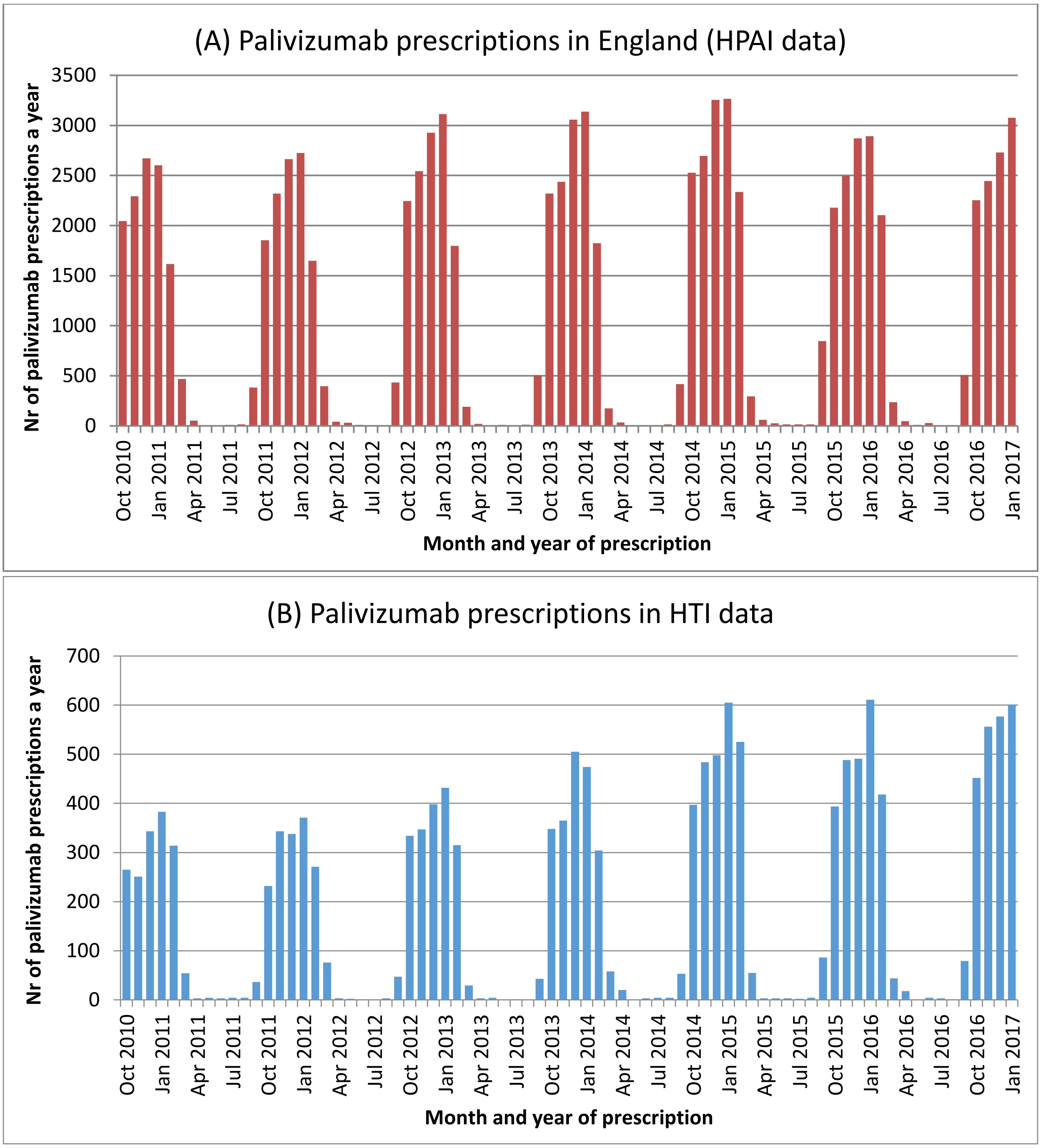


*HPAI: Hospital Pharmacy Audit Index, HTI=Hospital Treatment insights,*

##### 1.3.2 Summarising information about palivizumab prescription records

We removed palivizumab records dispensed between May and August. We were left with 14,757 records for 4,268 individuals (Appendix Figure 4).

Information about palivizumab dispensing was summarised for each child for each RSV season (referred to as child-seasons). There were 4,583 child-seasons for the 4,268 children in the HTI data. We removed child-seasons which did not link to a HES birth record and palivizumab prescriptions after child's first RSV season of life (*n*=688). The remaining 3,599 child-seasons were linked to the cohort of potentially eligible infants.

To investigate possible off-label prescribing, we applied the same exclusion criteria for dispensing records as for the cohort of eligible infants – we removed child-seasons for children who did not have full hospital admission history, died before the start of RSV season or were not under care of HTI-hospital and we selected one infant per multiple birth. We then separated prescribing in infants who were eligible, who were possibly eligible but had missing gestational age, and in infants who were not eligible.

*Appendix Figure 4 – Flow chart illustrating data cleaning for palivizumab records*

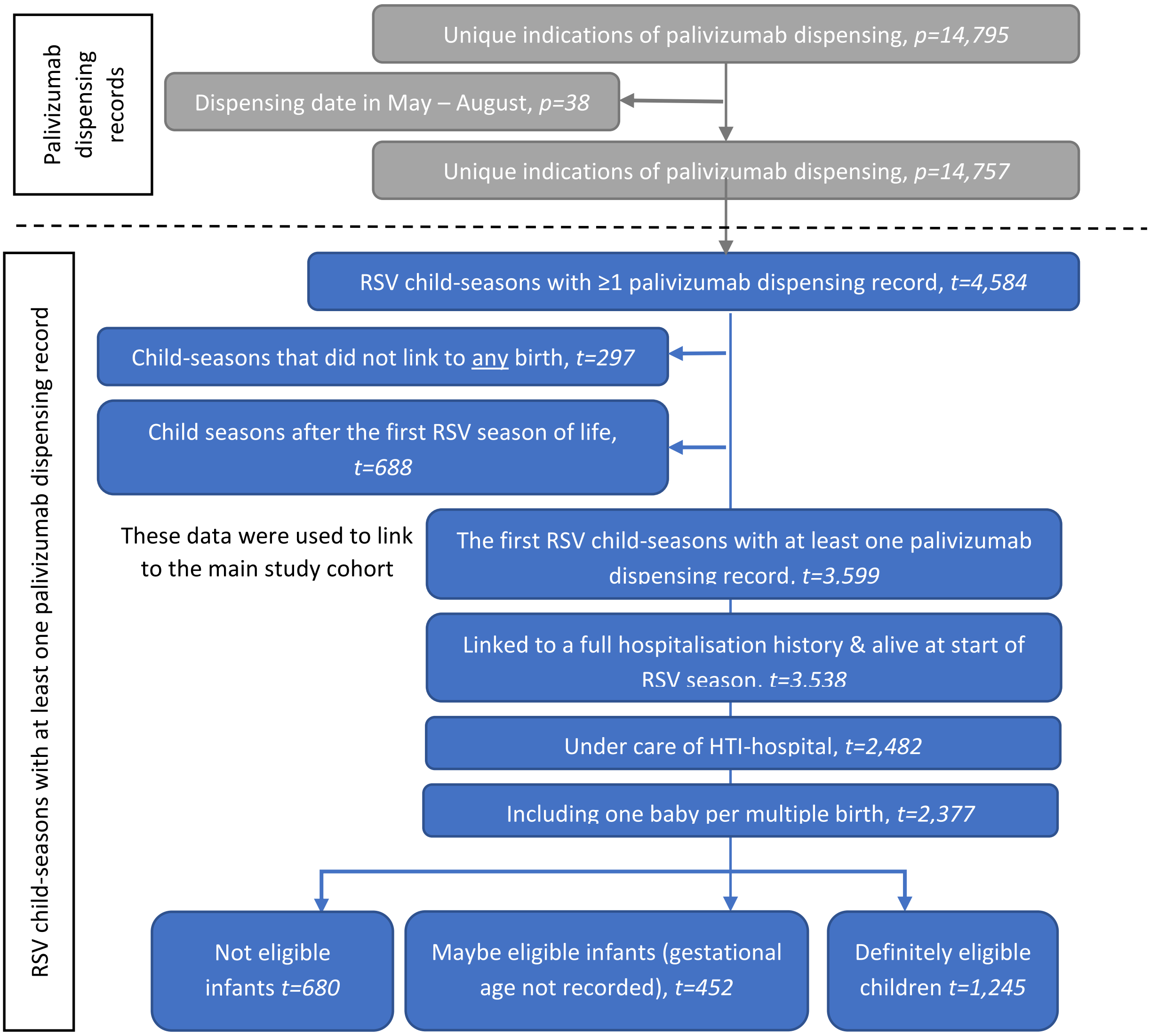


*HES=Hospital Episode Statistics, HTI=Hospital Treatment Insights, RSV=respiratory syncytial virus*

### 2 Appendix 2 – additional results

#### 2.1 Secondary analyses

*Appendix Table 4 – Characteristics of children who were treated with palivizumab*

| | Definitely eligible | Maybe eligible - missing gestational age | Not eligible |
|---|---|---|---|
| **Total (% of all)** | 1245 (52%) | 452 (19%) | 680 (29%) |
| **Age at start of RSV season** | | | |
| born during season | 203 (16%) | 78 (17%) | 100 (15%) |
| <1.5 months | 260 (21%) | 88 (19%) | 36 (5%) |
| 1.5-3 months | 261 (21%) | 93 (21%) | 158 (23%) |
| 3-6 months | 349 (28%) | 128 (28%) | 240 (35%) |
| 6-9 months | 172 (14%) | 65 (14%) | 131 (19%) |
| 9-12 months | *NA* | *NA* | 15 (2%) |
| **Eligibility conditions*** | | | |
| Heart condition | 882 (71%) | 212 (47%) | 207 (30%) |
| Lung condition | 1039 (83%) | 343 (76%) | 118 (17%) |
| **Gestational age (weeks)** | | | |
| <26 | 265 (21%) | | 28 (4%) |
| 26-27 | 322 (26%) | | 66 (10%) |
| 28-29 | 177 (14%) | | 111 (16%) |
| 30-31 | 57 (5%) | | 48 (7%) |
| 32-36 | 24 (2%) | | 77 (11%) |
| 37+ | 63 (5%) | | 194 (29%) |
| *Missing* | 337 (27%) | | 156 (23%) |
| **Birthweight (g)** | | | |
| <1000 | 554 (44%) | 52 (12%) | 113 (17%) |
| 1000-1499 | 256 (21%) | 46 (10%) | 130 (19%) |
| 1500-2499 | 63 (5%) | 28 (6%) | 96 (14%) |
| 2500+ | 62 (5%) | 21 (5%) | 199 (29%) |
| *Missing* | 310 (25%) | 305 (67%) | 142 (21%) |
| **Presence of other types of comorbidities** | | | |
| None | 142 (11%) | 144 (32%) | 240 (35%) |
| Respiratory | 363 (29%) | 106 (23%) | 124 (18%) |
| Cardiovascular | 229 (18%) | 76 (17%) | 125 (18%) |
| Neurological/sensory | 901 (72%) | 100 (22%) | 99 (15%) |
| Musculoskeletal/skin | 136 (11%) | 20 (4%) | 14 (2%) |
| Metabolic/endocrine/digestive/renal/genitourinary | 333 (27%) | 78 (17%) | 94 (14%) |
| Congenital anomalies | 303 (24%) | 90 (20%) | 180 (26%) |
| Cancers/blood | 167 (13%) | 54 (12%) | 67 (10%) |
| Chronic infections | 25 (2%) | <10 | <10 |
| Other chronic conditions | 194 (16%) | 68 (15%) | 113 (17%) |
| **Sex** | | | |
| Female | 551 (44%) | 188 (42%) | 301 (44%) |
| Male | 694 (56%) | 264 (58%) | 379 (56%) |
| **RSV season** | | | |
| 2010 | 129 (10%) | 46 (10%) | 69 (10%) |
| 2011 | 140 (11%) | 52 (12%) | 72 (11%) |
| 2012 | 182 (15%) | 58 (13%) | 98 (14%) |
| 2013 | 177 (14%) | 82 (18%) | 86 (13%) |
| 2014 | 219 (18%) | 86 (19%) | 137 (20%) |
| 2015 | 200 (16%) | 64 (14%) | 93 (14%) |
| 2016 | 198 (16%) | 64 (14%) | 125 (18%) |

*NA=not applicable, RSV=respiratory syncytial virus*

**Note that these categories are not mutually exclusive and column total will not add up to 100%*

#### 2.2 Sensitivity analyses

*Appendix Table 5 – Baseline characteristics of children who were eligible and had complete information on all risk factors, overall and by treatment status (strict definition)*

| | All eligible children | | Complete case cohort | |
|---|---|---|---|---|
| | N (%) of children in the cohort | N (%) treated | N (%) of children in the cohort | N (%) treated |
| **Total** | 1801 | 751 (42%) | 1588 | 673 (42%) |
| **Conditions related to eligibility*** | | | | |
| Congenital heart disease | 1271 (71%) | 510 (40%) | 1126 (71%) | 453 (40%) |
| Chronic lung disease | 1311 (73%) | 666 (51%) | 1153 (73%) | 598 (52%) |
| Severe combined immunodeficiency | 20 (1%) | 10 (50%) | 15 (1%) | <10 |
| 2 or more of above | 800 (44%) | 435 (54%) | 705 (44%) | 384 (54%) |
| **Gestational age (weeks)** | | | | |
| <26 | 451 (25%) | 234 (52%) | 409 (26%) | 213 (52%) |
| 26-27 | 584 (32%) | 276 (47%) | 527 (33%) | 254 (48%) |
| 28-29 | 413 (23%) | 149 (36%) | 373 (23%) | 133 (36%) |
| 30+ | 307 (17%) | 78 (25%) | 279 (18%) | 73 (26%) |
| *Missing* | 46 (3%) | 14 (30%) | | |
| **Birth weight (g)** | | | | |
| <1000 | 861 (48%) | 426 (49%) | 842 (53%) | 419 (50%) |
| 1000-1499 | 591 (33%) | 209 (35%) | 571 (36%) | 204 (36%) |
| 1500-2499 | 141 (8%) | 34 (24%) | 129 (8%) | 32 (25%) |
| 2500+ | 53 (3%) | 19 (36%) | 46 (3%) | 18 (39%) |
| *Missing* | 155 (9%) | 63 (41%) | | |
| **Sex** | | | | |
| Female | 812 (45%) | 329 (41%) | 715 (45%) | 295 (41%) |
| Male | 989 (55%) | 422 (43%) | 873 (55%) | 378 (43%) |
| **IMD Quintile** | | | | |
| Q1: least deprived 20% | 217 (12%) | 75 (35%) | 197 (12%) | 68 (35%) |
| Q2 | 292 (16%) | 120 (41%) | 270 (17%) | 110 (41%) |
| Q3 | 396 (22%) | 182 (46%) | 351 (22%) | 165 (47%) |
| Q4 | 377 (21%) | 150 (40%) | 336 (21%) | 130 (39%) |
| Q5: most deprived 20% | 471 (26%) | 213 (45%) | 434 (27%) | 200 (46%) |
| *Missing* | 48 (3%) | 11 (23%) | | |
| **Age at start of RSV season** | | | | |
| born during season | 711 (39%) | 156 (22%) | 624 (39%) | 145 (23%) |
| <1.5 months | 421 (23%) | 201 (48%) | 367 (23%) | 178 (49%) |
| 1.5-2 months | 323 (18%) | 188 (58%) | 291 (18%) | 167 (57%) |
| 3-5 months | 346 (19%) | 206 (60%) | 306 (19%) | 183 (60%) |
| **Presence of other types of comorbidities**** | | | | |
| No | 465 (26%) | 139 (30%) | 401 (25%) | 119 (30%) |
| Yes | 1336 (74%) | 612 (46%) | 1187 (75%) | 554 (47%) |
| **RSV season** | | | | |
| 2010/11 | 210 (12%) | 82 (39%) | 168 (11%) | 63 (38%) |
| 2011/12 | 260 (14%) | 90 (35%) | 230 (14%) | 82 (36%) |

| | | | | |
|---|---|---|---|---|
| 2012/13 | 264 (15%) | 114 (43%) | 242 (15%) | 105 (43%) |
| 2013/14 | 281 (16%) | 102 (36%) | 255 (16%) | 94 (37%) |
| 2014/15 | 281 (16%) | 129 (46%) | 250 (16%) | 116 (46%) |
| 2015/16 | 268 (15%) | 124 (46%) | 237 (15%) | 113 (48%) |
| 2016/17 | 237 (13%) | 110 (46%) | 206 (13%) | 100 (49%) |
| **Multiple birth** | | | | |
| Yes | 374 (21%) | 147 (39%) | 325 (20%) | 126 (39%) |
| No | 1427 (79%) | 604 (42%) | 1263 (80%) | 547 (43%) |

*IMD=Index of Multiple Deprivation, RSV=respiratory syncytial virus;*
**Note that these categories are not mutually exclusive and column total will not add up to 100%;*
*** These included congenital anomalies, cancer/blood disorders, chronic infections, metabolic/ endocrine/ digestive/ renal/ genitourinary conditions, musculoskeletal/skin conditions, neurological/sensory conditions or other non-specific chronic conditions)*

*Appendix Table 6 – Results from logistic regression models based on strict eligibility criteria*

| | Fixed effects model | Random effects model |
|---|---|---|
| **Complexity (SCID, CHD, CLD)** | | |
| 1 condition | 1 | 1 |
| 2 or more conditions | 2.04 (1.61, 2.60) | 2.16 (1.67, 2.80) |
| **Gestational age (weeks)** | | |
| <26 | 1.47 (1.00, 2.16) | 1.57 (1.04, 2.37) |
| 26-27 | 1.57 (1.10, 2.24) | 1.68 (1.15, 2.45) |
| 28-29 | 1.32 (0.91, 1.91) | 1.38 (0.94, 2.05) |
| 30+ | 1 | 1 |
| **Other chronic conditions** | | |
| No | 1 | 1 |
| Yes | 1.30 (1.00, 1.68) | 1.33 (1.00, 1.75) |
| **Age at start of RSV season** | | |
| born during season | 1 | 1 |
| <1.5 months | 3.22 (2.41, 4.29) | 3.52 (2.58, 4.78) |
| 1.5-2 months | 4.20 (3.08, 5.74) | 5.21 (3.70, 7.33) |
| 3-5 months | 4.13 (2.99, 5.70) | 5.08 (3.58, 7.21) |
| **Sex** | | |
| Female | 1 | 1 |
| Male | 1.08 (0.87, 1.35) | 1.04 (0.82, 1.31) |
| **IMD Quintile** | | |
| Q1: least deprived 20% | 1 | 1 |
| Q2 | 1.21 (0.80, 1.83) | 1.31 (0.83, 2.06) |
| Q3 | 1.42 (0.96, 2.10) | 1.43 (0.92, 2.21) |
| Q4 | 1.22 (0.82, 1.81) | 1.22 (0.78, 1.90) |
| Q5: most deprived 20% | 1.70 (1.16, 2.48) | 1.69 (1.09, 2.60) |
| **RSV season** | | |
| 2010 | 1 | 1 |
| 2011 | 0.89 (0.57, 1.39) | 0.88 (0.55, 1.43) |
| 2012 | 1.31 (0.84, 2.02) | 1.21 (0.75, 1.94) |
| 2013 | 0.91 (0.59, 1.41) | 0.87 (0.54, 1.39) |
| 2014 | 1.48 (0.96, 2.29) | 1.60 (1.00, 2.57) |
| 2015 | 1.54 (0.99, 2.39) | 1.48 (0.92, 2.38) |
| 2016 | 1.43 (0.91, 2.24) | 1.41 (0.86, 2.29) |

*CHD=congenital heart disease, CLD=chronic lung disease, IMD=Index of Multiple Deprivation, RSV=respiratory syncytial virus, SCID=severe combined immunodeficiency syndrome*
